# Targeted Metagenomics Reveals Hidden Chickenpox Epidemic Amid Mpox Surveillance in Uganda

**DOI:** 10.64898/2025.12.08.25341815

**Authors:** Stephen Kanyerezi, Alisen Ayitewala, Jupiter Marina Kabahita, Hellen Rosette Oundo, Julius Seruyange, Wilson Tenywa, Godwin Tusabe, Stacy Were, Moses Murungi, Martha Nabukyu, Valeria. Z. Nakintu, Caroline Makoha, Ivan Sserwadda, Harris Onywera, Collins Tanui, Ibrahim Mugerwa, Atek Kagirita, Benard Lubwama, Eyiru Micheal Rogers, David Patrick Kateete, Morgan Otita, Samuel Giduddu, Daudi Jjingo, Andrew Nsawotebba, Gerald Mboowa, Aloysious Ssemaganda, Susan Nabadda, Sofonias K Tessema, Isaac Ssewanyana

## Abstract

In regions where both Mpox virus (MPXV) and varicella zoster virus (VZV) are co-circulating, overlapping clinical manifestations can complicate clinical diagnosis. During the MPXV outbreak declared in Uganda on July, 2024, symptomatic suspected cases tested PCR negative for Mpox. To determine the cause of symptoms, we employed metagenomic sequencing with a targeted Viral Surveillance panel in 284 Mpox negative samples. VZV was identified as the predominant pathogen in 86% of MPXV-negative cases, suggesting a concurrent chickenpox surge. Using the VaricellaGen pipeline for variant calling, clade typing, and phylogeny, 118 (42%) samples that achieved ≥70% genome coverage were of clade 5 based on the single-nucleotide polymorphism (SNP) dataset. This data confirms co-circulation of chickenpox during the Mpox outbreak in Uganda. Our results underscore the need for laboratory confirmation of Mpox and the inclusion of VZV in the testing algorithm during the Mpox outbreak.

## Introduction

Mpox is an infection caused by the monkeypox virus (MPXV), a member of the *Orthopoxvirus* genus [1]. Mpox presents with a wide range of clinical symptoms such as fever, lymphadenopathy, and vesiculopustular lesions, similar to other exanthematous infections, particularly varicella-zoster virus (VZV), and has been reported in previous Mpox outbreaks [2]– [5]. While MPXV can be zoonotic, VZV is predominantly human-to-human transmission with no known animal reservoir [6]. The clinical overlap between these infections makes rapid and accurate diagnosis essential for effective outbreak response and management.

VZV is a common alphaherpesvirus with a double-stranded DNA genome known to cause primarily varicella (chickenpox) in children and adolescents and herpes zoster in older and or immunocompromised persons [7], [8]. VZV is highly contagious and is airborne with a genome of approximately 125 kb containing 71 open reading frames (ORFs) [9][10]. Currently, nine VZV clades are recognized: six established clades (1-6 and 9) and two provisional clades (VII and VIII) [11]. In Mpox-endemic regions, especially in low- and middle-income countries with low VZV vaccination coverage, chickenpox remains commonly reported [2]–[5].

Conventional laboratory methods for differentiating Mpox from VZV rely on polymerase chain reaction (PCR)-based assays, which are highly specific but limited to detecting only known targets. [12]. In outbreak settings, however, an overreliance on PCR may result in undetected alternative pathogens when cases test negative for the suspected virus [13]. For instance, a study during the 2014-2015 Ebola outbreak in Conakry, Guinea, found that among 2,362 suspected Ebola cases, 1,540 (65.2%) tested negative by PCR. Notably, 98 of these PCR-negative patients died, highlighting the potential presence of other severe illnesses that were initially overlooked due to the focus on Ebola [14]. In regions where both Mpox and VZV are co-circulating, overlapping clinical manifestations can complicate differential diagnosis, with Mpox frequently mistaken for VZV and vice versa [12], [15]. Recently, a study from Brazil deployed a multiplex PCR assay to diagnose co-infection of Mpox and VZV concurrently, but this has been limited in outbreak settings in Africa [4]. These findings underscore the need to consider alternative diagnoses and adopt comprehensive diagnostic strategies during outbreaks to avoid overlooking other clinically significant pathogens. Metagenomic sequencing offers an unbiased and powerful approach to pathogen detection, enabling the identification of viral agents beyond the initially suspected cause [16].

During this period, only 52% cases tested positive for Mpox despite presentation with symptoms. In order to understand other causes of symptoms among Mpox negatives, we utilized metagenomic sequencing.

## Methods

### Ethical consideration

Ethical approval was sought to utilize the Mpox outbreak investigation samples for this study from the Uganda National Health Laboratory Services Research and Ethics Committee (UNHLS-2025-133). This study was conducted in accordance with the principles outlined in the Declaration of Helsinki.

### Sample collection and PCR testing

During the MPXV outbreak declared in Uganda on July, 2024, 284 symptomatic suspected cases tested PCR negative for Mpox were included in this study. The samples included lesion, rash, oropharyngeal, or genital swabs in Viral Transport Media (VTM), as well as crusts collected in plain tubes - obtained as part of the public health emergency response to Mpox from affected individuals across 27 districts in the country. DNA was extracted using QIAamp DNA Kit (Qiagen, Hilden, Germany) and subsequently subjected to the Non-Variola Orthopoxvirus Real-Time PCR Primer and Probe Set (CDC, EUA) [17] in-house assay at the Central Emergency Response and Surveillance Laboratory (CERSL), Ministry of Health, Uganda. Briefly, amplification was conducted on the CFX96 Real Time PCR detection system BioRad thermocycler, with denaturation at 95 °C for 8 minutes, followed by 40 cycles, each consisting of two main steps i.e., a denaturation step at 95 °C for 5 seconds and an annealing and extension step at 60 °C for 30 seconds. PCR results were analyzed using CFX96 Real-Time PCR system (Bio-Rad Laboratories, Inc., California, USA) for Ct value interpretation and quality control.

### Metagenomic sequencing and taxonomic classification

High-quality DNA was extracted using the QIAamp DNA Kit (Qiagen, Hilden, Germany), normalized to a starting concentration of 50 ng, and prepared for metagenomic sequencing using a target enrichment approach with a pre-designed Illumina Viral Surveillance Panel (VSP). Paired-end sequencing was performed on the Illumina MiSeq platform at the Genomics Core Laboratory of the Central Public Health Laboratory. Raw sequencing reads were analyzed using KrakenUniq (v1.0.3) for taxonomic classification and identification of viral pathogens [18]. Using the developed VaricellaGen pipeline, reads were checked for quality, consensus genomes generated and only samples that attained genome coverage of 70% and above were subjected to downstream analysis. The same pipeline was used to assign clades to genomes and construct a phylogenetic tree. The resulting phylogenetic tree was visualized using the Interactive Tree of Life (iTOL v7) platform for intuitive exploration and annotation of clade relationships [19].

### VaricellaGen: Automated VZV genomic analysis pipeline development

To enhance the genomic characterization of VZV, we developed VaricellaGen (https://github.com/MicroBioGenoHub/VaricellaGen). The pipeline includes modules for quality control, variant calling, consensus genome generation, clade typing using a curated SNP dataset [20], and phylogenetic analysis with 1000 bootstraps and collapsing near to zero branch lengths. For the phylogenetic module, reference VZV sequences with known clade assignments were retrieved from NCBI and concatenated into a single FASTA file to serve as a background dataset [20]–[33]. The sequence JN704693.1 was designated as the outgroup. Sequence identifiers were standardized and annotated in the following format: GenBank accession, three-letter country code of sample origin, year of collection, and assigned clade.

### Variant calling and consensus genome assembly

The quality assessment of sequencing reads was designed to use FastQC (v0.12.1) and poor quality reads trimmed using Trim Galore (v0.6.10). The alignment of passed reads are aligned to the VZV reference genome (NC_001348.1) using bwa (v0.7.18-r1243-dirty) and samtools (v1.21), and variants are called using freebayes (v1.3.8) and BCFtools (v1.21). [34]–[38]. Process_gvcf from ARTIC was used to generate regions to be masked during consensus genome generation. [39]. To generate consensus sequences, VaricellaGen was designed to use BCFtools, and only samples that attained genome coverage of 70% and above were meant to be subjected to downstream analysis.

### Clade assignment and phylogenetic analysis

To classify the VZV strains, VaricellaGen used an integrated SNP-based clade typing module. For phylogenetic analysis, consensus genomes are concatenated and processed through VaricellaGen’s phylogeny module, which performs multiple sequence alignment using MAFFT (v7.525) and constructs a maximum-likelihood phylogenetic tree with IQ-TREE2 (v2.4.0) to assess the genetic relatedness of query samples to previously reported VZV strains [40], [41]. VaricellaGen uses reference VZV sequences with known clade assignments retrieved from NCBI and concatenated into a single FASTA file to serve as a background dataset. The sequence JN704693.1 was designated as the outgroup. Sequence identifiers were standardized and annotated in the following format: GenBank accession, three-letter country code of sample origin, year of collection, and assigned clade.

## Results

In this study, we analyzed 284 PCR-confirmed MPXV-negative cases from 27 districts (**Figure 1**). These included 151 (53.2%) males and 123 (43.3%) females, with a mean age of 14 years (range: 9 months - 70 years), who presented with Mpox-like symptoms, notably vesicular-pustular rash, fever, and lymphadenopathy (https://bit.ly/4iZeQDn). Gender information was unavailable for 10 (3.5%) isolates.

**Table 1:** Demographic and clinical characteristics of 284 PCR-confirmed MPXV-negative cases presenting with Mpox-like symptoms.

| Variable | Value |
| --- | --- |
| Total cases analyzed | 284 |
| MPXV status | PCR-confirmed MPXV-negative |
| Number of districts | 27 |
| Sex distribution | 151 (53.2%) males and 123 (43.3%) females. |
|  | Gender information unavailable for 10 cases (3.5%) |
| Mean age | 14 years |
| Age range | 9 months – 70 years |
| Clinical presentation | Vesicular-pustular rash, fever, lymphadenopathy |

**Figure 1.**
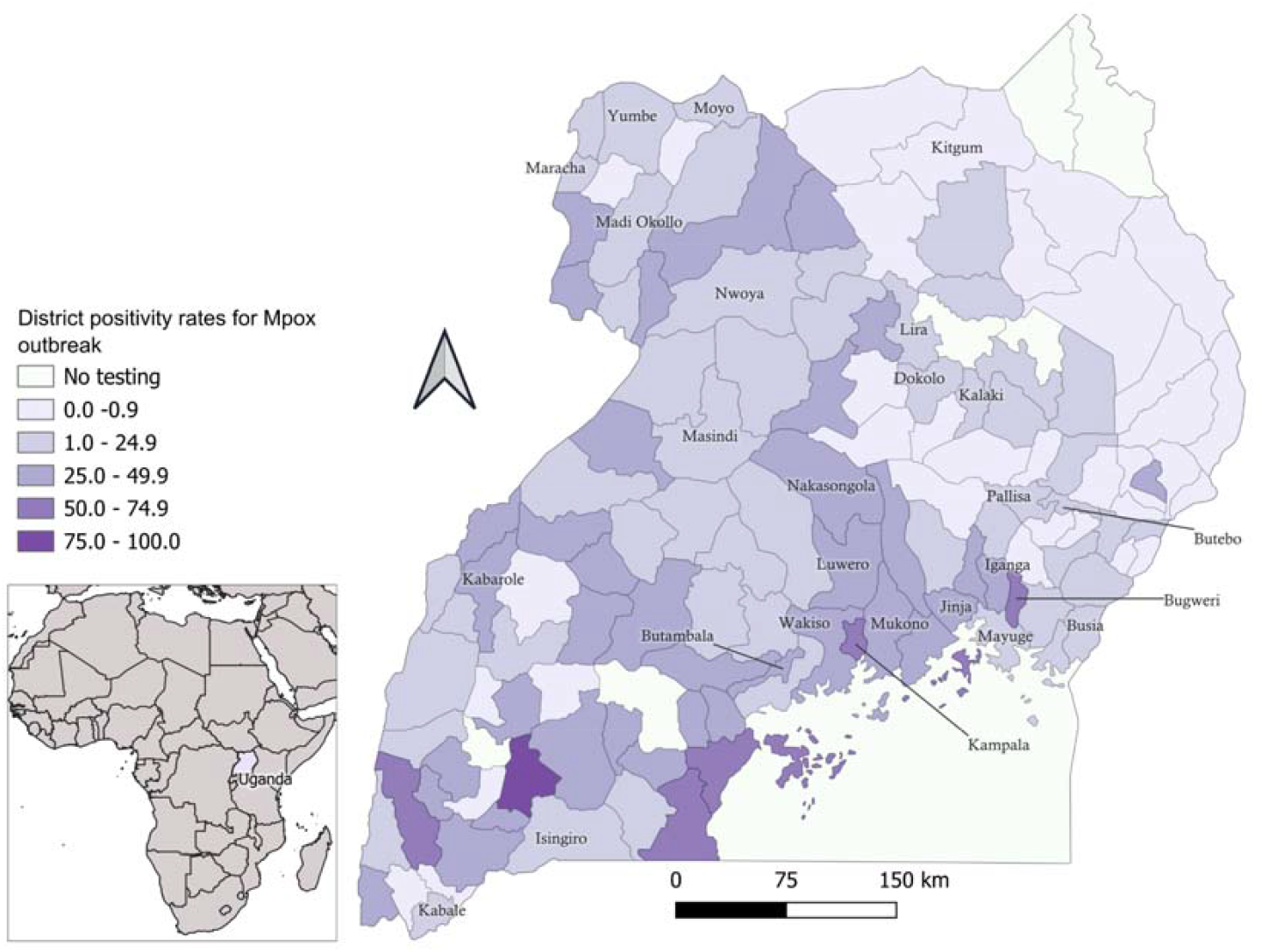
Geographic distribution of sample collection sites across Uganda. Map of Uganda showing Mpox outbreak and location of the 27 districts (labeled) from which MPXV-negative clinical samples were collected for metagenomic sequencing. Districts with no samples tested are not labeled.

PCR screening ruled out MPXV infection in all clinically suspected cases. Metagenomic sequencing, followed by taxonomic classification using KrakenUniq, identified VZV as the predominant viral pathogen. Specifically, 86% (243/284) of the samples had at least one read aligning to VZV, while 14% (41/284) showed no VZV reads.

### VaricellaGen: Automated VZV genomic analysis pipeline

To enhance the genomic characterization of VZV, we successfully developed VaricellaGen (https://github.com/MicroBioGenoHub/VaricellaGen), an automated pipeline for the genomic analysis of VZV. The pipeline integrates sequential modules for raw data quality control, variant calling, consensus genome generation, SNP-based clade typing [20], and phylogenetic reconstruction with 1000 bootstraps and collapsing near to zero branch lengths (**Figure 2**). Outputs include annotated consensus genomes, clade assignments, and a phylogenetic tree, providing a streamlined workflow from raw reads to interpretable genomic insights.

**Figure 2.**
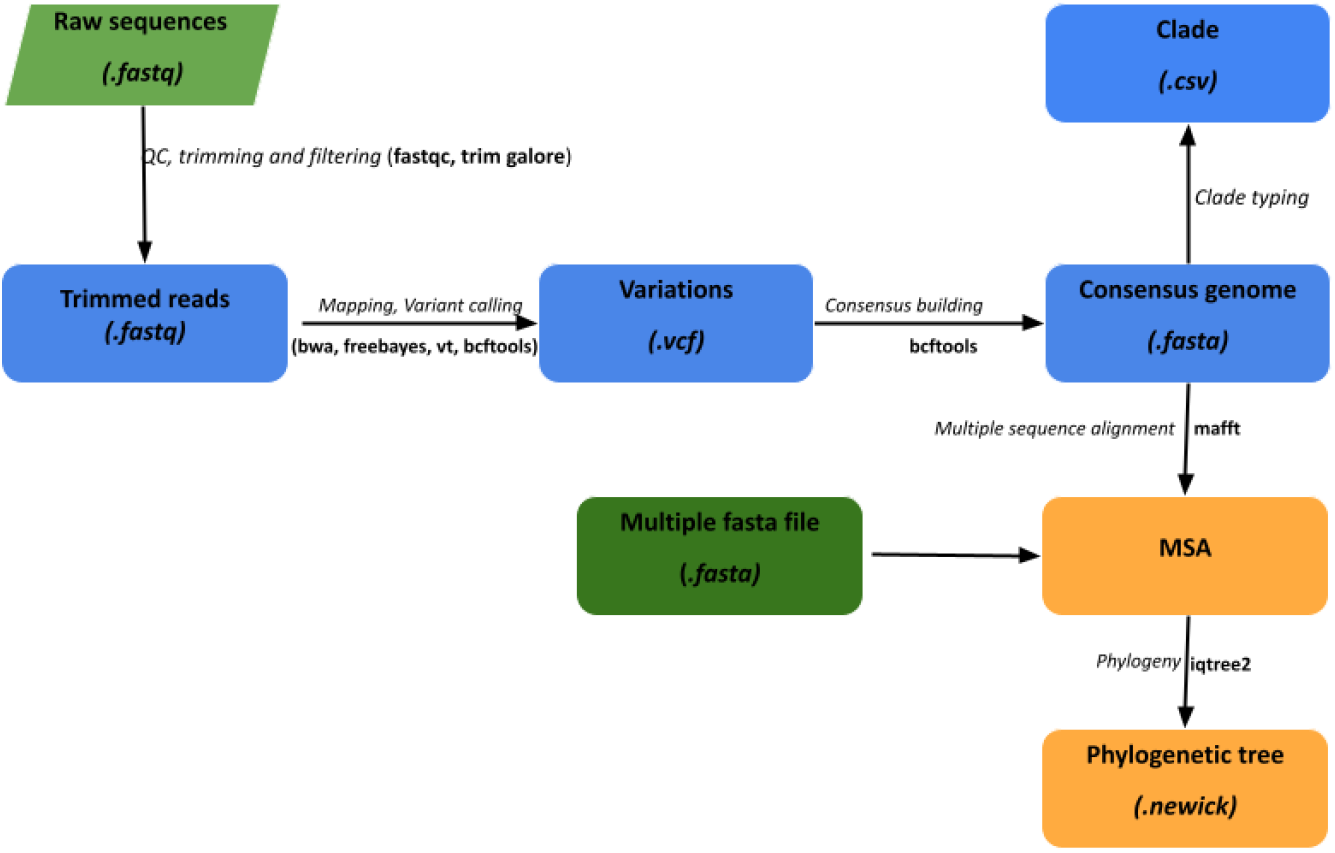
Flowchart of the VaricellaGen pipeline. The pipeline integrates sequential modules for raw data quality control, variant calling, consensus genome generation, clade typing (based on [20]), and phylogenetic reconstruction using a curated background dataset. Outputs include clade assignments, annotated consensus genomes, and phylogenetic trees for downstream interpretation.

### Clade assignment and phylogenetic insights

Of the 243 VZV-positive samples, 49% (118/243) achieved ≥70% genome coverage using the VaricellaGen pipeline. SNP-based clade typing classified all successfully sequenced samples as VZV clade 5. This finding was corroborated by phylogenetic analysis, which revealed two distinct sub-clusters within clade 5: one cluster predominantly composed of sequences from Ugandan, India, United States, and Kenyan samples; and a second cluster comprising the remaining Ugandan sequences along with one sequence from India (**Figure 3**).

**Figure 3.**
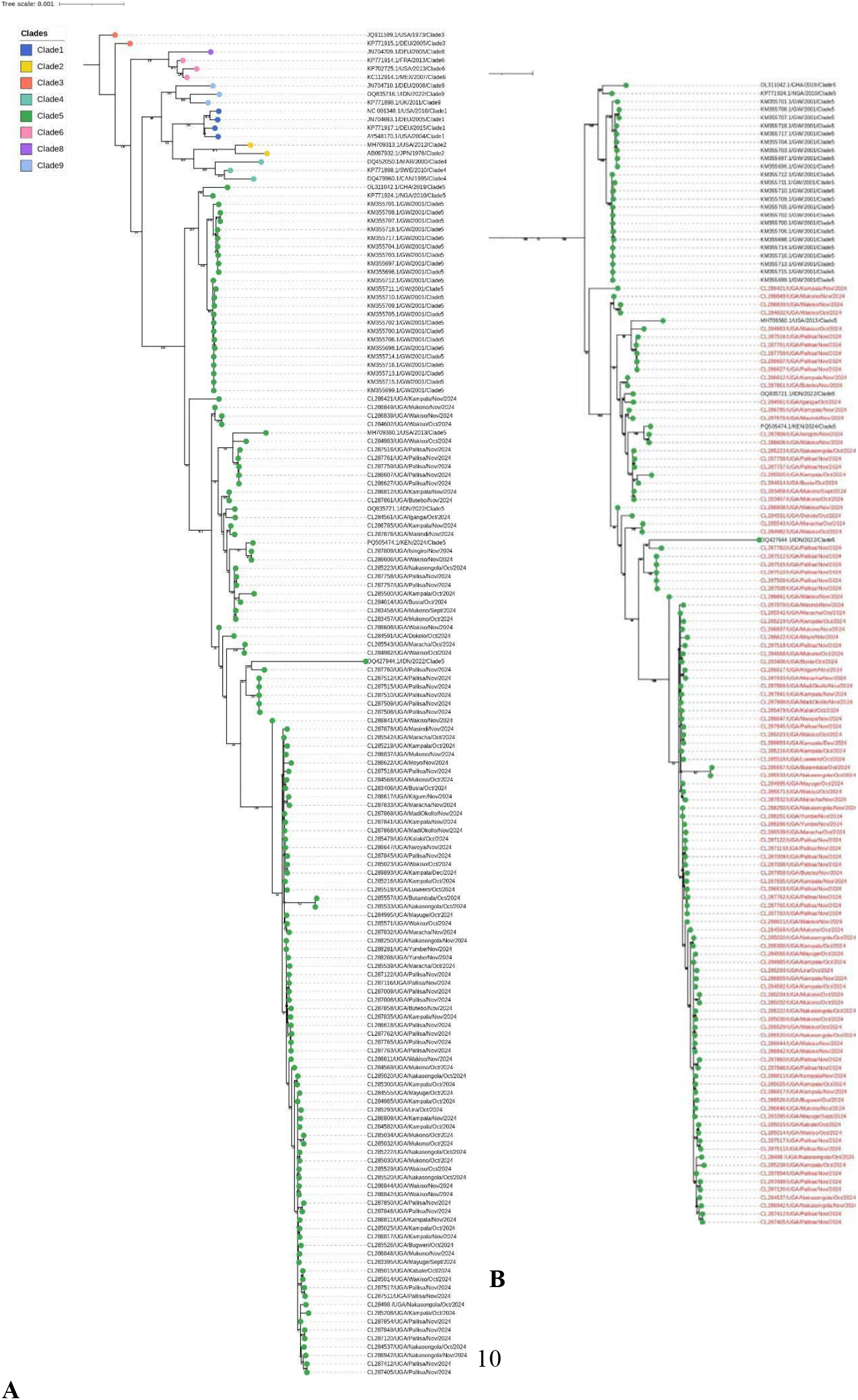
Phylogenetic analysis of Varicella Zoster Virus (VZV) genomes from Uganda. **A)** Maximum-likelihood phylogenetic tree of VZV genomes. Phylogenetic analysis of VZV-positive samples from Uganda (green) alongside reference sequences from GenBank representing known clades (Clades 1–9). **B)** All Ugandan samples (in red) clustered within Clade 5, forming two distinct sub-clusters: one grouping closely with sequences from India, the United States, and Kenya, and another clustering with only one sequence from India. The tree was constructed using IQ-TREE2 with MAFFT-based alignments; JN704693.1 (Clade 1) was used as the outgroup. Bootstrap values >30 are shown at key nodes. Tree visualization was performed using iTOL.

## Discussion

Genomic pathogen surveillance is critically important, especially during outbreaks with non-specific or overlapping clinical presentations. Here, we investigated cases that presented with Mpox-like signs but had MPXV infection ruled out by PCR. Metagenomic sequencing revealed VZV as the predominant etiological agent in a substantial proportion of cases, suggesting a concurrent and previously unrecognized surge in VZV infections during the Mpox outbreak in Uganda.

These findings are consistent with concurrent Mpox and VZV outbreaks reported in Burundi, Kenya, and the DRC [3], [4], [42], [43] and emphasize the challenge of differentiating co-circulating rash-causing viruses based on clinical signs alone [44], [45]. The similar skin manifestations of Mpox and VZV can complicate clinical diagnosis, especially in resource-limited settings with limited access to specific tests. This overlap underscores the necessity of comprehensive laboratory diagnostics, beyond single PCR assays, for accurate identification of the causative agent and effective public health responses during outbreaks.

Using a metagenomic sequencing approach, we identified VZV in 91% of PCR-confirmed MPXV-negative cases that were clinically suspected to be Mpox. Our newly developed VaricellaGen pipeline enabled detailed genomic analysis, showing that all samples with sufficient genome coverage belonged to VZV clade 5. Phylogenetic clustering with established clade 5 reference genomes further supported this classification and suggests that this clade is prevalent in Uganda, consistent with previous findings from other African countries [46], [47].

Our phylogenetic analysis of VZV clade 5 revealed two Ugandan sub-clusters. One mainly contained sequences from Uganda, India, US, and Kenya, suggesting regional lineage mixing or historical introductions. The other primarily included the remaining Ugandan sequences, clustering with one isolate from India, indicative of multiple introductions or diverse transmission chains within Uganda. This clustering pattern also points to travel-related spread or shared networks across distant regions. These findings highlight the importance of detailed genomic surveillance for both local and international transmission tracking. The observed sub-structuring within clade 5 strongly suggests the need for a standard naming system for VZV sub-clusters to improve genomic reporting and the precision of transmission analysis for future outbreak management.

The simultaneous detection of VZV and Mpox outbreaks highlights critical gaps in current outbreak response protocols, particularly in differential diagnosis and surveillance systems. Large-scale epidemics often focus on a single high-priority pathogen, inadvertently leading to missing concurrent infections, a pattern previously seen in influenza pandemics with underrecognized bacterial co-infections despite their significant contributions to morbidity and mortality [48]. These findings emphasize the need for integrated surveillance strategies with the ability to capture a wide spectrum of pathogens underscoring the need for multi-pathogen molecular diagnostics and genomic tools to avoid misclassification, especially in sub-Saharan Africa, where integrated disease surveillance systems have previously demonstrated the limitations of single-pathogen monitoring. [49]. A One Health approach, at the human, animal, and environment intersection further emphasizes the importance of monitoring multiple pathogens to detect emerging and re-emerging threats [50]. Our findings support the growing call for unbiased sequencing and comprehensive diagnostics in both routine surveillance and emergency responses.

Despite VZV being a DNA virus with a generally slow mutation rate [51], our analyzed sequences showed moderate genetic diversity (**Figure 3**), suggesting ongoing viral evolution or multiple introduction events. Since all sequences belonged to clade 5, the observed variation could indicate a within-clade origin reflective of probable local transmission patterns, accumulated mutations over time, or different points of origin.

Strengths of this study lie in the unbiased metagenomic sequencing, which allowed for a broad range of pathogen detection beyond Mpox, and the development of VaricellaGen, an automated pipeline for the rapid genomic characterization of DNA viruses (like VZV). By integrating quality control, variant calling, clade typing, and phylogenetics, VaricellaGen offers a scalable tool for outbreak genomic epidemiology. A key limitation of our study was that detailed genomic analyses only included samples with ≥70% genome coverage. While ensuring data quality, this may have excluded informative lower-quality samples, highlighting the need for robust sample collection and handling, as input quality impacts analysis [52]. While we identified clade 5 in our study, future research should explore if specific genomic features within this clade are associated with disease severity, transmission, or geographic spread.

Overall, our findings emphasize the crucial role of integrated genomic surveillance during outbreaks, especially when clinical presentation cannot be relied on to differentiate between circulating pathogens. The co-detection of VZV during Uganda’s Mpox outbreak shows the power of unbiased metagenomic sequencing and the need for robust diagnostic infrastructure to improve outbreak responses. This study further illustrates how genomic tools can aid in differential diagnosis, guide public health response, and reduce the risks of diagnostic ambiguity. The development of VaricellaGen offers a scalable, automated tool for detailed VZV genomic surveillance, providing a valuable resource for future outbreak investigations and public health decision-making

## Data availability

The source code and operation manual for VaricellaGen are available from GitHub under GNU GPL v3; (https://github.com/MicroBioGenoHub/VaricellaGen). The authors confirm that all supporting data, code, and protocols have been provided within the article. The genomic raw read files from this study are publicly available at the Sequence Read Archive (SRA) of the National Center for Biotechnology Information (NCBI) under the study BioProject ID: PRJNA1373217.

## Author contributions

Conceptualization: S.K., A.A., I.S., S.N., D.J., G.M., D.P.K, A.S., S.K.T., I.S. Formal analysis: S.K., A.A., J.M.K., C.M., I.S., G.M. Funding acquisition: S.N., I.S., S.T.K., H.O., C.T., N.A., A.A., A.S., S.K, I.M, A.K. Methodology: S.K., A.A., J.M.K., H.R.O., J.S., W.T., G.T., S.W., M.M., M.N., V.Z.N., C.M., I.S., G.M., A.S, I.M, B.L, E.M.R, M.O, S.G. Validation: S.K., A.A., B.A.K., H.O., C.T., N.A., V.Z.N., N.A., G.M., A.S., S.K.T., I.S. Visualization: S.K., I.S., J.M.K. Software: S.K., J.M.K., I.S., G.M. Writing – original draft: S.K., A.A., B.A.K., V.Z.N., I.S., H.O., C.T., N.A., S.N., D.J., G.M., A.A., S.K.T., I.S. Writing – review and editing: S.K., A.A., B.A.K., V.Z.N., I.S., H.O., C.T., N.A., S.N., D.J., G.M., A.A., S.K.T., I.S, I.M, B.L, E.M.R, M.O, S.G, A.K, D.P.K.

## Acknowledgement

We acknowledge the Ministry of Health Uganda and the Central Public Health Laboratories (CPHL) for supporting sample collection, diagnostic testing, and genomic sequencing during the Mpox outbreak response. We are grateful to the Genomics Core Facility at CPHL for sequencing support and to the field epidemiology and surveillance teams for their commitment to timely case investigation and sample coordination.

We also thank the Africa Centers for Disease Control and Prevention (Africa CDC) for their technical support and continued investment in strengthening genomic surveillance capacity in Uganda.

This work was supported by public health emergency response funds from the Government of Uganda. We further appreciate the contributions of colleagues who provided internal feedback during the preparation of this manuscript.

## References

[1] G. A. Shchelkunova and S. N. Shchelkunov, “Smallpox, Monkeypox and Other Human Orthopoxvirus Infections,” Viruses, vol. 15, no. 1, p. 103, Dec. 2022, doi:10.3390/v15010103.

[2] N. A. Hoff et al., “Varicella Coinfection in Patients with Active Monkeypox in the Democratic Republic of the Congo,” EcoHealth, vol. 14, no. 3, pp. 564–574, Sept. 2017, doi:10.1007/s10393-017-1266-5.

[3] C. M. Hughes et al., “A Tale of Two Viruses: Coinfections of Monkeypox and Varicella Zoster Virus in the Democratic Republic of Congo,” Am. J. Trop. Med. Hyg., vol. 104, no. 2, pp. 604–611, Feb. 2021, doi:10.4269/ajtmh.20-0589.

[4] P. R. Martins-Filho et al., “First reports of monkeypox and varicella-zoster virus coinfection in the global human monkeypox outbreak in 2022,” Travel Med. Infect. Dis., vol. 51, p. 102510, 2023, doi:10.1016/j.tmaid.2022.102510.

[5] A. Tayachew et al., “Genomic evidence of varicella-zoster virus among Mpox-suspected cases in Ethiopia during the 2022 Mpox multi-country outbreak,” Sci. Rep., Nov. 2025, doi:10.1038/s41598-025-29116-w.

[6] D. B. Di Giulio and P. B. Eckburg, “Human monkeypox: an emerging zoonosis,” Lancet Infect. Dis., vol. 4, no. 1, pp. 15–25, Jan. 2004, doi:10.1016/S1473-3099(03)00856-9.

[7] A. A. Gershon et al., “Varicella zoster virus infection,” Nat. Rev. Dis. Primer, vol. 1, p. 15016, July 2015, doi:10.1038/nrdp.2015.16.

[8] M. Wharton, “The epidemiology of varicella-zoster virus infections,” Infect. Dis. Clin. North Am., vol. 10, no. 3, pp. 571–581, Sept. 1996, doi:10.1016/s0891-5520(05)70313-5.

[9] A. A. Gershon, “Varicella zoster vaccines and their implications for development of HSV vaccines,” Virology, vol. 435, no. 1, pp. 29–36, Jan. 2013, doi:10.1016/j.virol.2012.10.006.

[10] J. Breuer, “Molecular Genetic Insights Into Varicella Zoster Virus (VZV), the vOka Vaccine Strain, and the Pathogenesis of Latency and Reactivation,” J. Infect. Dis., vol. 218, no. Suppl_2, pp. S75–S80, Sept. 2018, doi:10.1093/infdis/jiy279.

[11] V. T. Chow, G. A. Tipples, and C. Grose, “Bioinformatics of varicella-zoster virus: Single nucleotide polymorphisms define clades and attenuated vaccine genotypes,” Infect. Genet. Evol. J. Mol. Epidemiol. Evol. Genet. Infect. Dis., vol. 18, pp. 351–356, Aug. 2013, doi:10.1016/j.meegid.2012.11.008.

[12] A. K. Balingit et al., “Clinical presentation and molecular diagnosis of a possible Mpox virus and Varicella zoster virus co-infection in an adult immunocompetent Filipino: a case report,” Front. Public Health, vol. 12, p. 1387636, Nov. 2024, doi:10.3389/fpubh.2024.1387636.

[13] C. Mortier et al., “How to distinguish mpox from its mimickers: An observational retrospective cohort study,” J. Med. Virol., vol. 95, no. 10, p. e29147, 2023, doi:10.1002/jmv.29147.

[14] B. Ingelbeen et al., “Mortality among PCR negative admitted Ebola suspects during the 2014/15 outbreak in Conakry, Guinea: A retrospective cohort study,” PLoS ONE, vol. 12, no. 6, p. e0180070, June 2017, doi:10.1371/journal.pone.0180070.

[15] N. A. Hoff et al., “Varicella Coinfection in Patients with Active Monkeypox in the Democratic Republic of the Congo,” EcoHealth, vol. 14, no. 3, pp. 564–574, Sept. 2017, doi:10.1007/s10393-017-1266-5.

[16] W. Gu, S. Miller, and C. Y. Chiu, “Clinical Metagenomic Next-Generation Sequencing for Pathogen Detection,” Annu. Rev. Pathol., vol. 14, pp. 319–338, Jan. 2019, doi:10.1146/annurev-pathmechdis-012418-012751.

[17] CDC, “Non-variola Orthopoxvirus Real-time PCR Primer and Probe Set - EUA.” 2024.

[18] F. P. Breitwieser, D. N. Baker, and S. L. Salzberg, “KrakenUniq: confident and fast metagenomics classification using unique k-mer counts,” Genome Biol., vol. 19, no. 1, p. 198, Nov. 2018, doi:10.1186/s13059-018-1568-0.

[19] I. Letunic and P. Bork, “Interactive Tree Of Life (iTOL): an online tool for phylogenetic tree display and annotation,” Bioinforma. Oxf. Engl., vol. 23, no. 1, pp. 127–128, Jan. 2007, doi:10.1093/bioinformatics/btl529.

[20] N. J. Jensen et al., “Revisiting the genotyping scheme for varicella-zoster viruses based on whole-genome comparisons,” J. Gen. Virol., vol. 98, no. 6, pp. 1434–1438, June 2017, doi:10.1099/jgv.0.000772.

[21] “Human alphaherpesvirus 3 isolate KE_23082024, complete genome.” Nov. 24, 2024. Accessed: Apr. 15, 2025. [Online]. Available: http://www.ncbi.nlm.nih.gov/nuccore/PQ505474.1

[22] Y. Gomi, H. Sunamachi, Y. Mori, K. Nagaike, M. Takahashi, and K. Yamanishi, “Comparison of the Complete DNA Sequences of the Oka Varicella Vaccine and Its Parental Virus,” J. Virol., vol. 76, no. 22, pp. 11447–11459, Nov. 2002, doi:10.1128/JVI.76.22.11447-11459.2002.

[23] R. A. Santos, J. A. Padilla, C. Hatfield, and C. Grose, “Antigenic variation of varicella zoster virus Fc receptor gE: loss of a major B cell epitope in the ectodomain,” Virology, vol. 249, no. 1, pp. 21–31, Sept. 1998, doi:10.1006/viro.1998.9313.

[24] “Complete-Genome Phylogenetic Approach to Varicella-Zoster Virus Evolution: Genetic Divergence and Evidence for Recombination,” CoLab. Accessed: Apr. 15, 2025. [Online]. Available: https://colab.ws/articles/10.1128%2Fjvi.00835-06

[25] G. A. Peters et al., “A full-genome phylogenetic analysis of varicella-zoster virus reveals a novel origin of replication-based genotyping scheme and evidence of recombination between major circulating clades,” J. Virol., vol. 80, no. 19, pp. 9850–9860, Oct. 2006, doi:10.1128/JVI.00715-06.

[26] “Human herpesvirus 3 strain VZVi/Philadelphia.USA/73/V[3], partial genome.” May 16, 2012. Accessed: Apr. 15, 2025. [Online]. Available: http://www.ncbi.nlm.nih.gov/nuccore/JQ911599.1

[27] F. Garcés-Ayala et al., “Full-Genome Sequence of a Novel Varicella-Zoster Virus Clade Isolated in Mexico,” Genome Announc., vol. 3, no. 4, pp. e00752–15, July 2015, doi:10.1128/genomeA.00752-15.

[28] D. P. Depledge et al., “Evolution of Cocirculating Varicella-Zoster Virus Genotypes during a Chickenpox Outbreak in Guinea-Bissau,” J. Virol., vol. 88, no. 24, pp. 13936–13946, Dec. 2014, doi:10.1128/JVI.02337-14.

[29] P. Norberg et al., “Recombination of Globally Circulating Varicella-Zoster Virus,” J. Virol., vol. 89, no. 14, pp. 7133–7146, Apr. 2015, doi:10.1128/JVI.00437-15.

[30] N. J. Jensen et al., “Analysis of the reiteration regions (R1 to R5) of varicella-zoster virus,” Virology, vol. 546, pp. 38–50, July 2020, doi:10.1016/j.virol.2020.03.008.

[31] A. J. Davison and J. E. Scott, “The complete DNA sequence of varicella-zoster virus,” J. Gen. Virol., vol. 67 (Pt 9), pp. 1759–1816, Sept. 1986, doi:10.1099/0022-1317-67-9-1759.

[32] P. El-Duah et al., “Genetic characterization of varicella-zoster and HIV-1 viruses from the cerebrospinal fluid of a co-infected encephalitic patient, Ghana,” Virol. J., vol. 19, p. 122, July 2022, doi:10.1186/s12985-022-01854-7.

[33] A. Kumar et al., “First detection of Varicella Zoster Virus clade 9 cases in India during mpox surveillance,” Ann. Med., vol. 55, no. 2, p. 2253733, Dec. 2023, doi:10.1080/07853890.2023.2253733.

[34] P. Danecek et al., “Twelve years of SAMtools and BCFtools,” GigaScience, vol. 10, no. 2, p. giab008, Feb. 2021, doi:10.1093/gigascience/giab008.

[35] E. Garrison and G. Marth, “Haplotype-based variant detection from short-read sequencing,” July 20, 2012, arXiv: 1207.3907. doi:10.48550/arXiv.1207.3907.

[36] F. Krueger, FelixKrueger/TrimGalore. (Feb. 02, 2025). Perl. Accessed: Feb. 03, 2025. [Online]. Available: https://github.com/FelixKrueger/TrimGalore

[37] D. LaMar, “FastQC,” 2015, doi:https://qubeshub.org/resources/fastqc.

[38] H. Li and R. Durbin, “Fast and accurate short read alignment with Burrows–Wheeler transform,” Bioinformatics, vol. 25, no. 14, pp. 1754–1760, July 2009, doi:10.1093/bioinformatics/btp324.

[39] BCCDC-PHL, “BCCDC-PHL/mpxv-artic-nf.” Accessed: Apr. 08, 2025. [Online]. Available: https://github.com/BCCDC-PHL/mpxv-artic-nf

[40] B. Q. Minh et al., “IQ-TREE 2: New Models and Efficient Methods for Phylogenetic Inference in the Genomic Era,” Mol. Biol. Evol., vol. 37, no. 5, pp. 1530–1534, May 2020, doi:10.1093/molbev/msaa015.

[41] K. Katoh and D. M. Standley, “MAFFT Multiple Sequence Alignment Software Version 7: Improvements in Performance and Usability,” Mol. Biol. Evol., vol. 30, no. 4, pp. 772–780, Apr. 2013, doi:10.1093/molbev/mst010.

[42] C. Onyango et al., “High prevalence of varicella zoster virus infection among persons with suspect mpox cases during a mpox outbreak in Kenya, 2024,” Apr. 10, 2025, medRxiv. doi:10.1101/2025.04.08.25325502.

[43] N. Nzoyikorera et al., “Monkeypox Clade Ib virus introduction into Burundi: first findings, July to mid-August 2024,” Eurosurveillance, vol. 29, no. 42, p. 2400666, Oct. 2024, doi:10.2807/1560-7917.ES.2024.29.42.2400666.

[44] K. M. Derrick, J. W. Marson, S. Chakka, and E. R. Heilman, “Mpox virus and coinfections: An approach to rapid diagnosis,” J. Cutan. Pathol., vol. 50, no. 10, pp. 878–883, Oct. 2023, doi:10.1111/cup.14490.

[45] L. Guillén-Calvo et al., “Mpox, herpes, and enteroviruses: Differential diagnosis,” J. Med. Virol., vol. 96, no. 1, p. e29371, 2024, doi:10.1002/jmv.29371.

[46] A. Kumar et al., “First detection of Varicella Zoster Virus clade 9 cases in India during mpox surveillance,” Ann. Med., vol. 55, no. 2, p. 2253733, doi:10.1080/07853890.2023.2253733.

[47] P. Bryant, T. Yildirim, S. B. Griesemer, K. Shaw, D. Ehrbar, and K. St George, “Vaccine Strain and Wild-Type Clades of Varicella-Zoster Virus in Central Nervous System and Non-CNS Disease, New York State, 2004-2019,” J. Clin. Microbiol., vol. 60, no. 4, p. e0238121, Apr. 2022, doi:10.1128/jcm.02381-21.

[48] D. E. Morris, D. W. Cleary, and S. C. Clarke, “Secondary Bacterial Infections Associated with Influenza Pandemics,” Front. Microbiol., vol. 8, p. 1041, 2017, doi:10.3389/fmicb.2017.01041.

[49] I. R. Mremi, J. George, S. F. Rumisha, C. Sindato, S. I. Kimera, and L. E. G. Mboera, “Twenty years of integrated disease surveillance and response in Sub-Saharan Africa: challenges and opportunities for effective management of infectious disease epidemics,” One Health Outlook, vol. 3, no. 1, p. 22, Nov. 2021, doi:10.1186/s42522-021-00052-9.

[50] T. R. Kelly et al., “Implementing One Health approaches to confront emerging and reemerging zoonotic disease threats: lessons from PREDICT,” One Health Outlook, vol. 2, no. 1, p. 1, Jan. 2020, doi:10.1186/s42522-019-0007-9.

[51] G. A. Peters et al., “The Attenuated Genotype of Varicella-Zoster Virus Includes an ORF0 Transitional Stop Codon Mutation,” J. Virol., vol. 86, no. 19, pp. 10695–10703, Oct. 2012, doi:10.1128/jvi.01067-12.

[52] S. S. Ajay, S. C. J. Parker, H. Ozel Abaan, K. V. Fuentes Fajardo, and E. H. Margulies, “Accurate and comprehensive sequencing of personal genomes,” Genome Res., vol. 21, no. 9, pp. 1498–1505, Sept. 2011, doi:10.1101/gr.123638.111.

